# Frequent vs single active bouts differentially affect movement behavior and energy balance in adults with overweight/obesity

**DOI:** 10.64898/2026.04.14.26350871

**Authors:** Heloisa C. Santo Andre, Elisa Le Roux, Nathan P. De Jong, Patricia R. Smith, Andrew H. Lange, Carlos Mendez, Alexandre Zahariev, Melissa L. Mamele, Ginger Johnson, Zhaoxing Pan, Chantal Simon, Daniel H. Bessesen, Ana J. Pinto, Audrey Bergouignan

**Affiliations:** Division of Endocrinology, Metabolism and Diabetes, Anschutz Health and Wellness Center, Department of Medicine, School of Medicine, University of Colorado Anschutz Medical Campus, Aurora, CO, United States; Faculty of Applied Sciences, State University of Campinas, SP, Brazil; Institut Pluridisciplinaire Hubert Curien, Centre National de la Recherche Scientifique, Université de Strasbourg UMR7178, Strasbourg, France; Section of Endocrinology, Department of Pediatrics, School of Medicine, University of Colorado Anschutz Medical Campus, Aurora, CO, United States; CarMeN Laboratory, INSERM 1060, INRA 1397, University Claude Bernard Lyon 1, Human Nutrition Research Center Rhône-Alpes, Oullins, France

**Keywords:** energy expenditure, doubly labeled water, appetite, accelerometry, humans

## Abstract

**Objective:** To investigate the effects of breaking up prolonged sedentary behavior (SB) on daily movement behavior and energy balance in adults with overweight/obesity.

**Methods:** Thirty participants (16F/14M; 34.2±7.3y; 29.5±3.2kg/m^2^) were randomized to either BREAK (nine hourly 5-min brisk walking bouts) or a duration-matched intervention, ONE (45-min brisk walking), both performed 5 days/week for 6 weeks. Pre- and post-intervention, daily SB and physical activity (PA; accelerometry), body composition (doubly labeled water [DLW]), total daily energy expenditure (TDEE; DLW), appetite, and fasting leptin were measured. Linear-mixed effects models tested time effects and time-by-group interactions.

**Results:** Only BREAK reduced prolonged SB (-8%; interaction: p=0.043). Both groups shifted SB-PA composition toward greater moderate-to-vigorous PA with proportional reductions in SB and light PA (time: all p≤0.011), which were associated with increases in TDEE (+0.67 MJ/d; time: p=0.040). Body and fat mass increased in ONE only (interaction: p=0.061 and p=0.055). No differences were noted in energy intake, appetite, or leptin levels.

**Conclusions:** Spreading short PA bouts throughout the day increases MVPA and TDEE to the same extent as a traditional continuous PA bout. Future studies should investigate whether minor differences in body composition are driven by distinct behavioral/physiological compensations influenced by the daily pattern of PA/SB.

**STUDY IMPORTANCE QUESTIONS:**

1. **What is already known about this subject?**
  - Acutely, breaking up prolonged sedentary behavior (SB) with short bouts of physical activity (PA) increases energy expenditure and reduces food cravings compared to a single continuous PA bout.
  - Single continuous PA bouts have been associated with compensatory reductions in non-exercise activities (daily living activities) in some studies, which may attenuate increases in total daily energy expenditure (TDEE) and limit effects on body mass and adiposity.
2. **What are the new findings in your manuscript?**
  - Performing brisk walking either through frequent, short bouts spread across the day or as a single continuous bout over 6 weeks increases moderate-to-vigorous PA (MVPA) at the expense of SB and light PA and increases TDEE to a similar extent in adults with overweight or obesity.
  - However, only the frequent, short active breaks reduced time spent in prolonged SB (>60 min), an independent cardiometabolic health risk factor.
  - Despite no differences in energy intake, appetite, or plasma leptin concentration, the single continuous bouts were associated with a small, non-robust increase in body and fat mass, whereas these remained stable in the active breaks group, suggesting differential compensatory adaptations.
3. **How might your results change the direction of research or the focus of clinical practice?**
  - Promoting frequent, short bouts of PA throughout the day can improve daily movement and help meet current PA/SB guidelines to a similar extent as traditional PA strategies, while also reducing prolonged sedentary time.
  - This strategy may help limit compensatory responses sometimes observed in response to continuous MVPA bouts, offering a new tool to manage body weight.
  - However, differences in body composition outcomes were small and not robust, and future studies are needed to determine whether these patterns translate into meaningful long-term effects on energy balance and weight regulation.

## INTRODUCTION

Sedentary behavior (SB), characterized by low energy expenditure (≤1.5 METS), such as sitting, reclining, or lying down, is an independent risk factor for chronic conditions, including insulin resistance, type 2 diabetes, or obesity (1). In Westernized countries, objective assessments indicate that adults spend approximately 8-12 h/day sedentary (2, 3). Higher SB is associated with greater body mass index (BMI), waist circumference, total and trunk fat mass (FM) (4, 5).

Breaking up prolonged SB with short bouts of PA has been associated with reductions in body mass (BM) and waist circumference, independent of overall SB or MVPA levels (6). A meta-analysis including data from 33 studies conducted in adults with normal weight or overweight/obesity reported modest reductions in BM, waist circumference, and body fat percentage following SB reduction interventions (7).

BM loss reflects cumulative negative energy balance over time, i.e., when TDEE exceeds energy needs. Acutely, short bouts of low-intensity walking (1-to-5-min) increase energy expenditure compared with uninterrupted sitting and standing breaks to SB (8, 9). Our group showed that spreading moderate-intensity PA over 6 hours reduced pre-meal food cravings compared with a single 30-minute bout in healthy adults (10). Similarly, replacing sitting time (-21%) with standing reduced appetite and energy intake (EI) in office workers over 4 weeks (11). These findings suggest that breaking up SB may increase TDEE while decreasing EI, supporting a more favorable energy balance. However, no study has examined the impact of active breaks on both EI and TDEE over extended periods. Furthermore, it remains unclear whether the pattern of PA accumulation itself influences energy balance beyond the effect of increasing total MVPA.

The objective of this 6-week study was to compare the effects of breaking up SB with frequent, short PA bouts versus a traditional single PA bout matched for total active time (45 min/day, 5 d/week) on daily SB and PA, BM, body composition, and energy balance components. We hypothesized that spreading regular active breaks would more effectively reduce prolonged SB and limit spontaneous behavioral adaptation (i.e., decreases in light PA - LPA) than performing a single continuous bout, thereby promoting a more favorable energy balance.

## METHODS

### Study Design

This was a single-center, parallel-group, randomized controlled trial (RCT, ClinicalTrials.gov: NCT02998892; Figure 1). Potentially eligible participants were invited for a screening visit, which included a medical history and physical examination, a fasting blood draw, the International Physical Activity Questionnaire [IPAQ] to assess habitual PA, and a 5-day assessment of habitual step count using a pedometer. Eligible participants were randomized to either intervention: ONE or BREAK. Habitual SB/PA, TDEE, resting metabolic rate (RMR), appetite, and leptin concentration were measured at baseline (Pre) and at the end of the intervention (Post).

**Figure 1.**
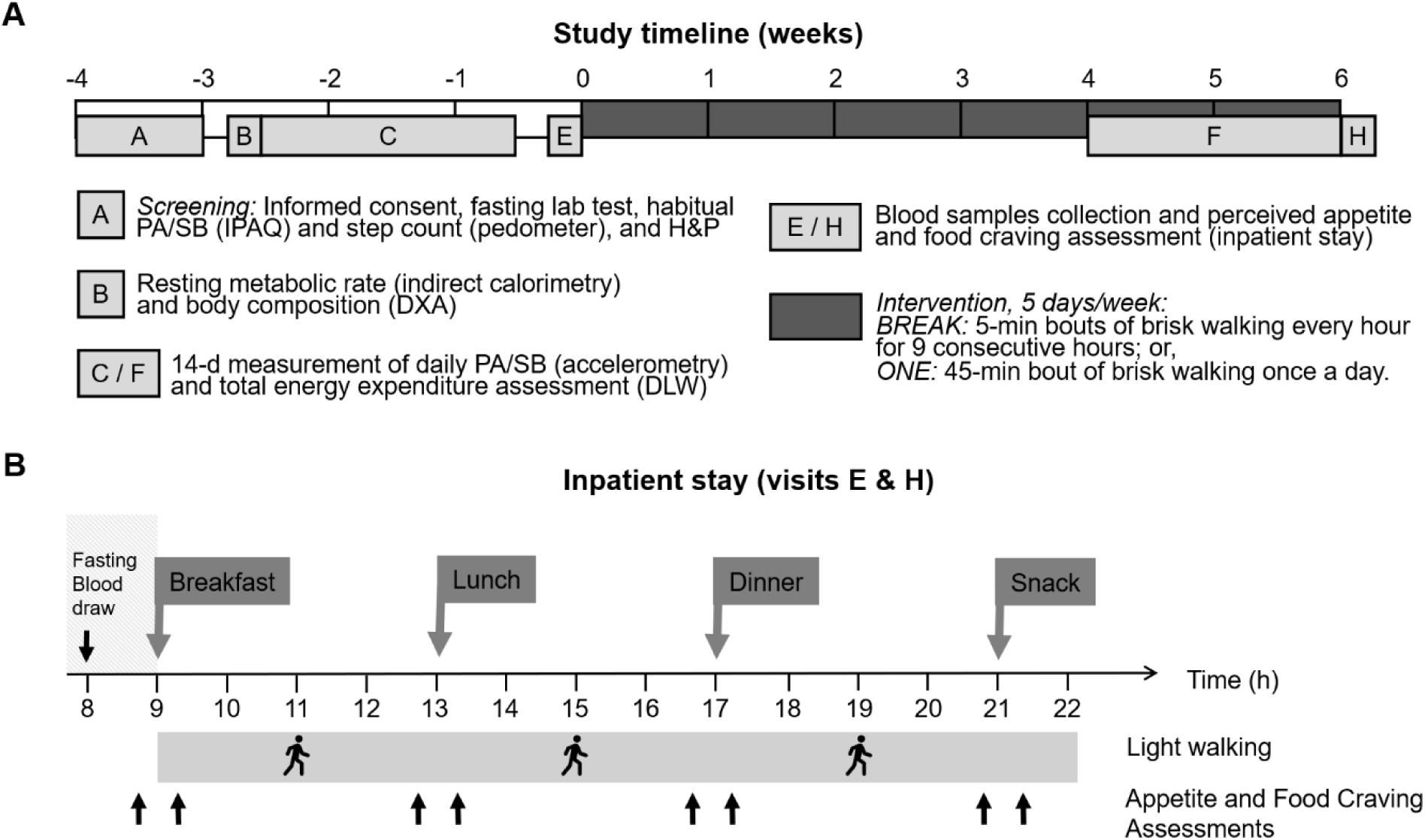
Study design overview. Abbreviations: IPAQ, International Physical Activity Questionnaire; H&P, health and physical examination; DXA, dual-energy X-ray absorptiometry; RMR, resting metabolic rate; PA, physical activity; SB, sedentary behavior; DLW, doubly labeled water.

### Ethics approval

This study was approved by the Colorado Multiple Institutional Review Board (COMIRB; protocol 16-1769) and conducted in accordance with the Declaration of Helsinki (12). Written informed consent was obtained from each participant.

### Participants

Eligible participants: 18–45 years, BMI 25–38 kg/m², weight stable for >6 months, sedentary (>6 h/day sitting), physically inactive (<150 min/week MVPA and/or <6,500 steps/day). Exclusion criteria included insulin resistance, diabetes, cardiovascular, renal, hepatic disease, uncontrolled hypertension, cancer, HIV, substance abuse (drugs, tobacco, or >40 g/day alcohol), psychiatric disorders, medications affecting lipid/energy metabolism, PA contraindications, and pregnancy/lactation/perimenopause/post menopause. Participants were recruited from May 2017 to October 2021 in the Denver metro area (Colorado).

### Intervention

Participants were randomized (1:1), stratified by sex, to either (1) ONE, moderate-intensity brisk walking for 45 min, 5 day/wk; or (2) BREAK, ten 5-min bouts of brisk walking every hour, 5 day/wk. In BREAK, ten bouts were prescribed rather than nine to account for the approximately 10% lower MVPA typically completed in free-living conditions (13), ensuring about 45 min/day of completed activity. All participants completed a treadmill familiarization to determine individualized moderate pace, starting at 2.0 mph with 0.3 mph increments every 2 minutes; the speed corresponding to a Borg rating of 6 (“somewhat hard”) out of 10 was used for the PA bouts. The study statistician generated the allocation sequence and placed it into numbered opaque envelopes. Outcome assessors and researchers who delivered the interventions were not blinded.

The intervention lasted 5.7±1.9 weeks. Adherence was monitored using a wrist-worn Fitbit® Charge 2 (San Francisco, CA, US), worn continuously throughout the intervention. Participants were considered adherent if > 80% of prescribed PA was achieved (i.e., ≥ 36 min/day of continuous PA in ONE and ≥ 7 bouts/day in BREAK; ≥ 4 d/wk for both). To support compliance, participants tracked progress via the Fitbit app and received regular motivational messages or individualized follow-ups. They were asked to maintain their habitual diet and avoid intentional weight loss.

### Sedentary behavior and physical activity

Participants wore an ActivPAL4 (right thigh; PALTechnologies, Glasgow, Scotland) and an ActiGraph GT3X+ (hip, right side; Ametris, Fort Walton Beach, FL) for 14 consecutive days at baseline and during the final two intervention weeks. The ActiGraph was removed during sleep and water-based activities. A daily log captured sleep and non-wear time. Valid data required ≥10 h/day on ≥4 days, including one weekend day (14, 15). Mean wear time for ActiGraph was 13.6±1.2 h/day, and mean waking time for activPAL was 15.7±1.1 h/day.

ActiGraph GT3X+ quantifies PA (inactivity, LPA, and MVPA). Data were cleaned (e.g., non-wear time identification and removal of invalid days), analyzed, and exported using ActiLife6 software, v. 6.11.9 (Ametris, Pensacola, Florida). Freedson cut points were used to estimate inactivity (<100 counts/min), LPA (≥100 to <1952 counts/min), and MVPA (≥1952 counts/min) (16). Inactivity refers mainly to SB, but includes standing or minimal movement.

ActivPAL4 assesses postural allocation (sitting, standing, and stepping). Data were cleaned (e.g., sleep time adjustment, removal of non-valid days), analyzed, and exported using PALanalysis software, v.7.2.32 (PAL Technology, Glasgow, UK). These metrics of SB were computed: total SB (time spent sitting, in seated transport, and secondary lying down), total breaks in SB (number of times a sitting/lying down event was followed by a standing or stepping event), and time in prolonged SB bouts ≥60 minutes. Total time spent standing and stepping, and daily step count were computed. Because sleeping time was removed from analysis, SB mainly corresponds to waking SB.

### Total daily energy expenditure and its components

TDEE was measured using doubly labeled water (DLW) method over 14 days, as previously described (17). Subjects ingested a premixed 2g/kg of estimated total body water (TBW) dose of DLW composed of 0.2 and 0.15 g/kg of 10% enriched H_2_ ^18^O and 99% enriched ^2^H_2_O (CIL, Andover, MA), respectively (18, 19). Urine samples were collected in duplicates every other day for the determination of ^2^H and ^18^O enrichment by isotope ratio mass spectrometry (Delta V Advantage IRMS). Details on equilibration and endpoint urine cleaning, mass spectrometry procedures, quality check controls, and calculations are published elsewhere (20). TDEE was calculated using Schoeller’s equation (21), with individual 24-hour respiratory quotients measured during the inpatient stay in a whole-room calorimeter at baseline and post-intervention. Activity energy expenditure (AEE) was calculated as AEE_(MJ/day)_ = TDEE_(MJ/day)_ – diet-induced thermogenesis (DIT)_(MJ/day)_ – RMR_(MJ/day)_. RMR was measured via indirect calorimetry with a metabolic cart (ParvoMedics TrueOne 2400, Salt Lake City, UT) for 20 minutes after an overnight fast, at rest and thermoneutrality (22), and DIT was assumed to be 10% of TDEE. PA level (PAL) was calculated as participants’ TDEE divided by their RMR.

### Body Composition

BM was measured using a calibrated scale. At baseline, body composition was measured by dual-energy X-ray absorptiometry (DXA; Hologic Delphi-W, Bedford, MA, USA) (23). Changes in body composition were estimated by isotopic dilution. Fat-free mass (FFM) from DLW was calculated by estimating a hydration coefficient of 73.2%, and FM was calculated as the difference between BM and FFM (18, 19). The agreement between the DXA and hydrometry data at baseline was 0.26 kg (-2.05 to 2.58) for FFM (r^2^=0.99, p<0.001).

### Metabolized Energy Intake

Metabolizable EI was calculated using the principle of energy balance (24, 25) as: EI_(MJ/day)_ = TDEE_(MJ/day)_ + ΔES_(MJ/day)_. Changes in energy store (ΔES) were calculated using the individual body composition change predicting model (26):

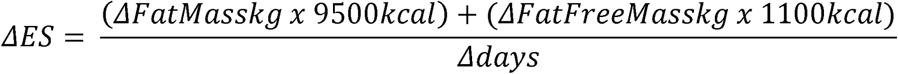

### Perceived appetite and food cravings

During pre and post 24-hour inpatient visits, participants received a standardized diet (55% carbohydrate, 30% fat, and 15% protein of EI) calculated to match their energy needs. Meals were provided at 0900, 1300, 1700, and 2100h (30%, 30%, 30%, and 10% of daily EI). Individual energy needs were estimated as RMR x activity factor, with RMR derived from the average of measured values (indirect calorimetry) and DXA-based estimates ([23.9 x FFM in kg] + 372) (13, 27).

Participants rated hunger and prospective food consumption before meals and fullness 1-hour post-meal, using a visual analog scale (VAS). The Food Craving Questionnaire – State (15 items; 1-5 scale) was completed before each meal (28). Total score reflects craving intensity; subscales (29) assessed desire to eat, anticipated reward, relief from negative states, loss of control, and physiological craving (hunger). Meal scores were averaged to obtain daily values.

### Fasting leptin concentrations

On the day of the 24-hour inpatient visits, an intravenous catheter was inserted into an antecubital vein for a blood draw. Plasma leptin concentrations were measured using an ultra-sensitive ELISA kit (Ultrasensitive Leptin ELISA Kit, ALPCO®; 22-LEPHUU-E01; intra-assay CV: 2.63% - 4.35%; inter-assay CV: 6.1% - 7.5%:) in the Beckman Coulter AU480 Chemistry Analyzer (Brea, CA).

### Statistical analysis

Participant characteristics were summarized using descriptive statistics.

The primary inferential focus was on the group-by-time interaction, expressed as the estimated mean difference in change between groups (EMD; ΔBREAK − ΔONE) with 95% confidence intervals (95%CI). When the interaction was not statistically significant, the main effect of time was examined and reported as the estimated time difference (ETD; Post − Pre) with 95%CI.

All dependent variables, except for step count, were analyzed using linear mixed models (LMMs) with group (BREAK vs ONE) and time (Pre vs Post) as fixed factors, and participant as a random factor, using a compound symmetry (CS) covariance structure.

Time-dependent variables for daily PA and SB were analyzed using a compositional approach, as previously described (30). Movement behaviors were expressed as 3-part compositions (ActiGraph: inactivity, LPA, and MVPA; activPAL: SB, standing, and stepping) and transformed using isometric log-ratio (ilr) coordinates. Two ilr coordinates were derived per system (e.g., ilr1 representing inactivity relative to LPA and MVPA, and ilr2 representing the balance between LPA and MVPA), and the ilr system was rotated to allow interpretation of each behavior individually. Only ilr1 is reported in the main text, as it captures the primary contrast of interest, whereas ilr2 estimates and stacked LMMs including both ilr coordinates and a balance factor were explored as sensitivity analyses to assess internal redistribution of behaviors.

Daily step count was analyzed using a generalized LMM with group, time and their interaction as fixed factors, participant as a random factor, a negative binomial distribution with log link, and wear time included as an offset to account for exposure and overdispersion.

Analyses were adjusted for sex, with additional covariates depending on the outcome: waking time for sit-to-stand transitions; BM for TDEE and its components; squared-height for BM, FM and FFM; BM and energy balance as measured during the 24hr test day in the whole-room calorimeter (difference between 24h TDEE measured from gas exchange and accurately measured EI (13)) for appetite and food cravings; and BM for fasting leptin. Additional analyses adjusted for sex only are presented in the supplemental material.

Analyses followed the intention-to-treat (ITT) principle, with missing data handled using LMMs via restricted maximum likelihood estimation under the assumption that data were missing completely at random or missing at random. The sample sizes of valid data for each outcome are described in **Supplementary Table 1,** along with the reasons for missing data.

LMMs residual diagnostics were assessed graphically (Q–Q plots and residuals versus fitted values) to evaluate normality and homoscedasticity of conditional residuals, and influential observations were examined using studentized residuals. For generalized LMM with a negative binomial distribution, model adequacy was assessed using appropriate residual and overdispersion diagnostics.

Variables were analyzed on their original scale unless a transformation was required to satisfy model assumptions. Appetite, food cravings, and fasting leptin were Blom-transformed due to non-normal distributions of model residuals.

Sensitivity analyses were conducted by excluding extreme observations and two participants who deviated from the protocol (i.e., one exceeded the prescribed PA dose, and another one unexpectedly traveled during Post data collection). Results from sensitivity analyses that differ from ITT are discussed.

Data analysis was performed using SAS 9.4 (SAS Institute Inc., Cary, NC, USA). The study statistician was not blinded to group allocation. Data are presented as mean ± standard deviation (SD), median (25th, 75th percentile), or least square means (LSMeans) with 95%CI, as appropriate. For compositional data, model estimates (LSMeans) obtained on ilr coordinates were back-transformed to the original scale for interpretation. Briefly, ilr coordinates were rescaled, exponentiated, and normalized to obtain compositional proportions expressed in min/day or percentages. Back-transformed ETD and EMD represent multiplicative changes over time and multiplicative between-group differences in change, respectively. Significance was set at p≤0.05 for main effects and p≤0.10 for interaction terms. No adjustment for multiple comparisons was applied.

## RESULTS

### Participants’ characteristics

The CONSORT diagram is shown in **Supplementary Figure 1**. Thirty participants were randomized to BREAK (n=8F/7M) or ONE (n=8F/7M) groups. Participants had a mean age of 34.2±7.3 years, BMI of 29.5±3.2 kg/m², and adiposity of 37.8±7.9%. Baseline HOMA-IR was 1.5±0.6, indicating participants did not have insulin resistance (**Table 1)**.

**Table 1.** Participant characteristics.

|  | **BREAK**<br>**(n=15)** | **ONE**<br>**(n=15)** |
| --- | --- | --- |
| **Sex** |  |  |
| Female | 8 [53.3%] | 8 [53.3%] |
| Male | 7 [46.7%] | 7 [46.7%] |
| **Age (years)** | 33.7 ± 7.6 | 34.7 ± 7.3 |
| **Race (n [%])** |  |  |
| White | 10 [66.7%] | 10 [66.7%] |
| Black | 4 [26.7%] | 2 [13.3%] |
| Other | 1 [6.7%] | 3 [20.0%] |
| **Ethnicity (n [%])** |  |  |
| Hispanic | 2 [13.3%] | 6 [40.0%] |
| Non-Hispanic | 13 [86.7%] | 9 [60.0%] |
| **Body mass (kg)** | 87.0 ± 15.5 | 84.2 ± 10.1 |
| **Height (cm)** | 170.0 ± 8.6 | 169.8 ± 9.4 |
| **BMI (kg/m<sup>2</sup>)** | 29.9 ± 4.2 | 29.1 ± 1.9 |
| **Fat-free Mass (kg)** | 53.0 ± 10.0 | 52.0 ± 10.3 |
| **Fat Mass (kg)** | 33.1 ± 11.0 | 31.1 ± 6.3 |
| **Fat Mass (%)** | 38.0 ± 8.6 | 37.7 ± 7.5 |
| **HOMA-IR** | 1.6 ± 0.5 | 1.5 ± 0.7 |
| **RMR (kcal)** | 1 584.3 ± 246.9 | 1 592.6 ± 232.0 |
| **5-d average daily steps** | 4 639 ± 1 026 | 4 867 ± 1 185 |
| **Self-reported sitting (h/day)** | 10.6 ± 2.8 | 10.5 ± 2.8 |

Data presented as mean ± SD or n [%]. Abbreviations: BMI, body mass index; MET, metabolic equivalent of task; MVPA, moderate-to-vigorous physical activity; RMR, resting metabolic rate.

### Both interventions increased MVPA and reduced sedentariness, but only BREAK reduced prolonged sitting

**Table 2** summarizes PA/SB outcomes. Participants’ adherence to the intervention was high with no significant difference between BREAK and ONE (94.3±15.0% vs 96.6±9.4%; p=0.606). A decrease in the percentage of total SB spent in prolonged bouts (>60min) was observed in BREAK (within-group change: -7.8; 95%CI: -13.5 to -2.2), whereas no difference was observed in ONE (within-group change: 0.6, 95%CI: -5.2 to 6.4; interaction: p=0.043). However, none of the interventions modified sit-to-stand transitions (time: p=0.701, interaction: p=0.288). No other time-by-group interactions were noted for any PA/SB outcomes, including the compositional changes (interaction: all p≥0.34).

**Table 2.** Habitual sedentary behavior and physical activity level.

| Outcomes | BREAK (n=15) |  | ONE (n=15) |  | ETD<br>(95%CI) | EMD<br>(95%CI) | group<br>effect, p | time<br>effect, p | interaction<br>effect, p |
| --- | --- | --- | --- | --- | --- | --- | --- | --- | --- |
|  | Pre | Post | Pre | Post |  |  |  |  |  |
| Intervention |  |  |  |  |  |  |  |  |  |
| Adherence (%) | - | 94.3 ±15.0 | - | 96.6 ± 9.4 | - | - | 0.606 | - | - |
| Intervention duration (wk.) | - | 5.7 ± 2.1 | - | 5.7 ± 1.8 | - | - | 0.963 | - | - |
| ActivPAL<sup>†</sup> |  |  |  |  |  |  |  |  |  |
| Total SB (min/day) | 631.2 (565.9, 696.6) | 602.8 (542.0, 663.7) | 644.7 (581.6, 707.8) | 596.5 (536.5, 656.5) | 0.80 (0.67, 0.94) | 1.03 (0.74, 1.45) | 0.453 | 0.011 | 0.842 |
| Compositional mean (min/day [%]) | 637.5 (69.6%) | 609.4 (65.9%) | 654.7 (68.3%) | 607.4 (63.1%) |  |  |  |  |  |
| Standing time (min/day) | 205.2 (164.4, 246.1) | 211.6 (174.6, 248.7) | 212.7 (175.2, 250.2) | 235.7 (192.0, 279.3) | 0.96 (0.87, 1.06) | 0.91 (0.75, 1.11) | 0.678 | 0.357 | 0.341 |
| Compositional mean (min/day [%]) | 200.1 (21.9%) | 207.8 (22.5%) | 206.4 (21.5%) | 228.3 (23.7%) |  |  |  |  |  |
| Stepping time (min/day) | 79.3 (64.2, 94.4) | 109.8 (87.4, 132.2) | 101.5 (80.9, 122.1) | 131.0 (103.8, 158.2) | 1.31 (1.16, 1.48) | 1.07 (0.84, 1.36) | 0.096 | <0.001 | 0.581 |
| Compositional mean (min/day [%]) | 78.1 (8.5%) | 107.2 (11.6%) | 97.8 (10.2%) | 127.1 (13.2%) |  |  |  |  |  |
| Prolonged SB ≥60 min (% total SB)<sup>‡</sup> | 34.3 (28.8, 39.8) | 26.5 (21.0, 32.0) | 29.5 (24.0, 35.0) | 30.1 (24.4, 35.7) | -3.6 (-7.7, 0.4) | -8.4 (-16.5, -0.3) | 0.855 | 0.079 | 0.043 |
| Sit-to-stand transitions (number/day)<sup>‡</sup> | 44.4 (39.5, 49.3) | 45.4 (40.5, 50.3) | 50.7 (45.8, 55.5) | 48.5 (43.5, 53.5) | -0.6 (-3.5, 2.4) | 3.1 (-2.8, 9.0) | 0.144 | 0.701 | 0.288 |
| Step count (number/day)<sup>‡</sup> | 6477 (5427, 7729) | 9559 (8003, 11419) | 7490 (6269, 8949) | 10611 (8839, 12739) | 1.45 (1.25, 1.68) | 1.04 (0.78, 1.40) | 0.221 | <0.001 | 0.778 |
| ActiGraph<sup>†</sup> |  |  |  |  |  |  |  |  |  |
| Inactivity time (min/d) | 577.0 (546.1, 608.0) | 567.5 (527.7, 607.3) | 542.5 (484.2, 600.7) | 546.6 (480.6, 612.6) | 0.68 (0.52, 0.89) | 0.87 (0.51, 1.49) | 0.669 | 0.007 | 0.600 |
| Compositional mean (min/day [%]) | 584.7 (72.3%) | 580.3 (70.4%) | 547.4 (67.6%) | 555.3 (67.5%) |  |  |  |  |  |
| Light-intensity PA (min/day) | 211.0 (183.8, 238.2) | 206.3 (173.4, 239.2) | 249.0 (199.2, 298.7) | 242.6 (185.3, 299.8) | 0.65 (0.53, 0.79) | 0.95 (0.64, 1.42) | 0.117 | <0.001 | 0.803 |
| Compositional mean (min/day [%]) | 208.3 (25.8%) | 205.4 (24.9%) | 247.8 (30.6%) | 236.1 (28.7%) |  |  |  |  |  |
| Moderate-to-vigorous PA (min/day) | 20.9 (13.2, 28.6) | 50.8 (25.7, 75.9) | 18.3 (12.0, 24.6) | 33.5 (23.1, 43.9) | 2.24 (1.50, 3.33) | 1.24 (0.56, 2.74) | 0.461 | <0.001 | 0.589 |
| Compositional mean (min/day [%]) | 15.7 (1.9%) | 38.8 (4.7%) | 14.6 (1.8%) | 31.3 (3.8%) |  |  |  |  |  |

Daily step count increased in both groups by 45% (ETD fold change: 1.45, 95%CI: 1.25 to 1.67; time: p<0.001). Both interventions increased time spent in MVPA (ETD: 2.24, 95%CI: 1.50 to 3.33; time: p<0.001), with absolute estimates rising from 15.7 to 38.8 min/day in BREAK and from 14.6 to 31.3 min/day in ONE. This greater dominance of MVPA was accompanied by proportional reductions in inactive time (ETD: 0.68, 95%CI: 0.52 to 0.89; time: p=0.007) and time spent in LPA (ETD: 0.65, 95%CI: 0.53 to 0.79; time: p<0.001) within the composition, although absolute changes were modest. Stepping time also increased (ETD: 1.31, 95%CI: 1.16 to 1.48; time: p<0.001), with absolute estimates rising from 78.1 to 107.2 min/day in BREAK and from 97.8 to 127.1 min/day in ONE. This greater dominance of stepping was accompanied by a proportional reduction in total SB time (ETD: 0.80, 95%CI: 0.67 to 0.94; time: p=0.011), but not standing time (ETD: 0.96, 95%CI: 0.87 to 1.06; time: p=0.357). Results from ilr2 and stacked models confirmed that the patterns of change observed in ilr1 were consistent with the internal redistribution of behaviors, and conclusions were unchanged. Raw ilr1 and ilr2 coordinates are reported in **Supplementary Table 2**. Sensitivity analyses yielded the same results as ITT analyses.

### BREAK and ONE significantly increased TDEE to a similar extent

No group × time interactions were observed for energy outcomes (all p > 0.74). There was a significant overall increase in TDEE_adjBM&sex_ (ETD: +0.67 MJ/day, 95%CI: 0.03 to 1.31; time effect: p=0.040, interaction: p=0.652; **Figure 2A**). That was driven by slight, non-significant increases in AEE_adjBM&sex_ in both groups (ETD: +0.51 MJ/day, 95%CI: -0.04 to 1.05; time: p=0.069, interaction: p=0.537; **Figure 2B**); RMR_adjBM&sex_ was not significantly modified in neither group (ETD: +0.10 MJ/day, 95%CI: -0.06 to 0.26; time: p=0.200; interaction: p=0.736; **Figure 2C**). Although not significant, PAL tended to increase in both groups (time: p=0.065; interaction: p=0.504; **Figure 2D**). Sensitivity analyses yielded similar results. Data not adjusted for weight are available in **Supplementary Table 3**.

**Figure 2.**
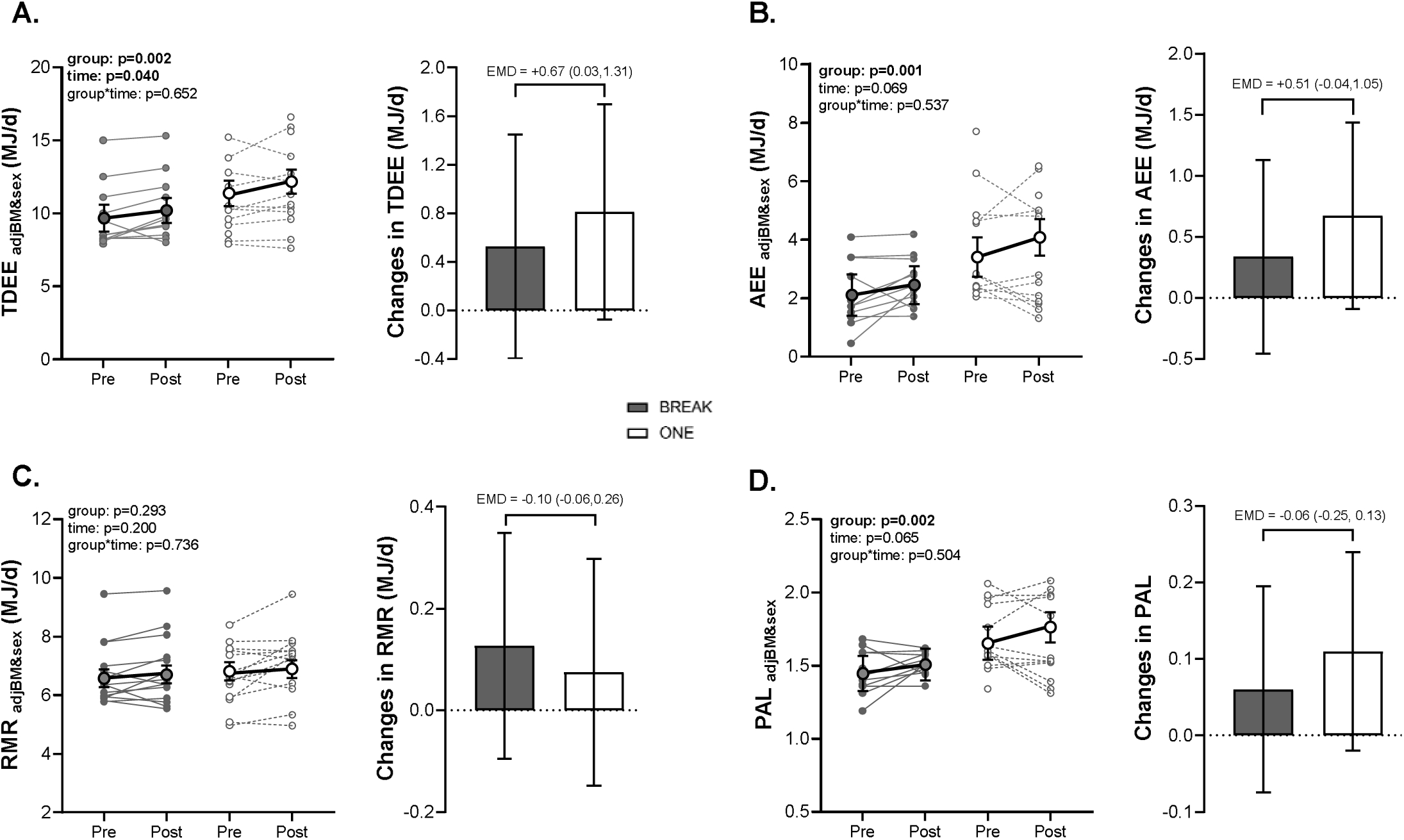
Energy expenditure and its components changes during the protocol. (A) Changes in total daily energy expenditure (TDEE). (B) Changes in activity energy expenditure (AEE). (C) Changes in resting metabolic rate (RMR). (D) Changes in physical activity level (PAL). Data are presented as individual values and LSMeans (95%CI) for graphs on the left, and within-group changes (95%CI) along with EMD (95%CI) for graphs on the right. Estimated mean difference (EMD) refers to differences between changes in BREAK and in ONE (ΔBREAK - ΔONE). LSMeans and EMD are derived from linear mixed models (LMMs) that included group, time, and group-by-time as fixed effects and study participants as random effects, with CS as the covariance structure; LMMs were adjusted for sex and body mass. For TDEE, AEE, and PAL, the sample size is n=13 for BREAK and n=14 for ONE. For RMR, the sample sizes are n=15 in both groups. Abbreviations: EMD: estimated mean difference; MVPA, moderate-to-vigorous physical activity.

### Minor increase in body mass and fat mass following ONE but not BREAK

A significant group x time interaction was observed for BM (p=0.061; **Figure 3A**), reflecting an increase in ONE (+ 1.5 kg; 95%CI: 0.35 to 2.58), but no change in BREAK (-0.1 kg; 95%CI: - 1.2 to 1.1). A similar interaction was observed for FM (ONE: +2.4 kg; 95%CI: 0.5 to 4.3; BREAK: -0.2 kg; 95%CI: -1.2 to 1.6; interaction: p=0.055; **Figure 3B**), as FFM was not modified (time: p=0.653, interaction: p=0.727; **Figure 3C**).

**Figure 3.**
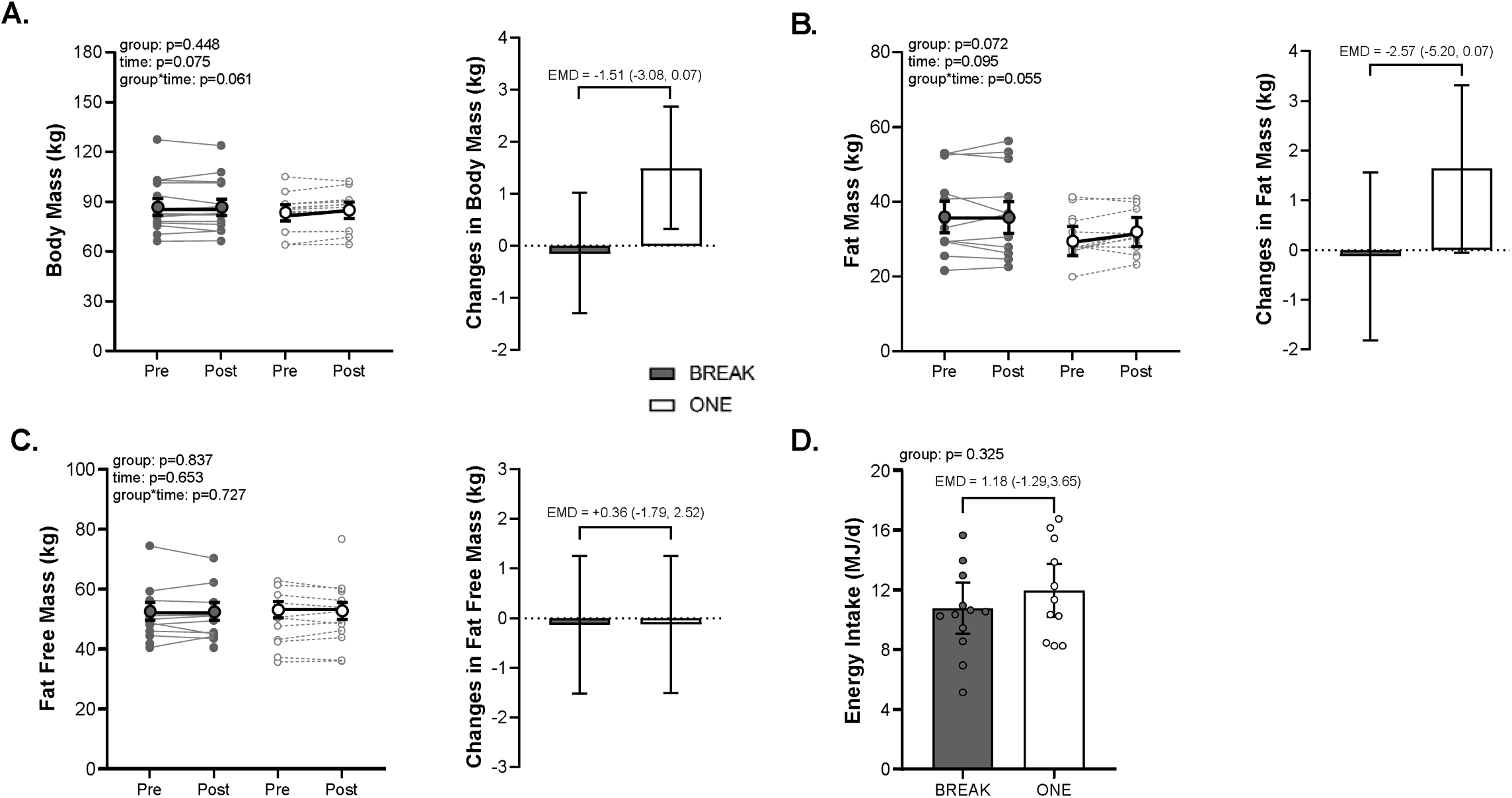
Body mass and composition changes during the protocol and energy intake during the interventions. (A) Changes in body mass. (B) Changes in fat mass. (C) Changes in fat-free mass. (D) Energy intake during the interventions. Data are presented as individual values and LSMeans (95%CI) for graphs on the left, and within-group changes (95%CI) along with EMD (95%CI) for graphs on the right. Estimated mean difference (EMD) refers to differences between changes in BREAK and in ONE (ΔBREAK - ΔONE). LSMeans and EMD are derived from linear mixed models (LMMs) that included group, time, and group-by-time as fixed effects and study participants as random effects with CS as the covariance structure; LMMs were adjusted for squared height and sex for body mass and body composition, and for sex only for estimated energy intake. For body mass, fat-free mass and fat mass, the sample size is n=12 in both groups. For energy intake, the sample size is n=10 for BREAK and n=9 for ONE.

Sensitivity analyses excluding the participant who exceeded the prescribed PA dose in BREAK attenuated these interactions (BM: p=0.116; FM: p=0.106) and indicated a significant overall increase over time in BM (+0.9 kg; 95%CI: -0.2 to 1.6; time p=0.018) and FM (+1.4 kg; 95%CI: -0.2 to 2.6; time p=0.026), still driven by the ONE group.

### No effect on self-perceived appetite, food cravings, leptin concentrations, or estimated EI

Estimated daily EI during the intervention period did not differ between groups (EGD: +1.18 MJ/day, 95%CI: -1.29 to 3.65; p=0.325; **Figure 3D**).

Appetite- and food cravings-related scores remained unchanged (all p>0.17; **Table 3**), as well as fasting leptin concentrations (time: p=0.112; interaction: p=0.928; **Table 3**). Appetite, food craving, and leptin data adjusted only for sex are available in **Supplementary Table 3**.

**Table 3.**
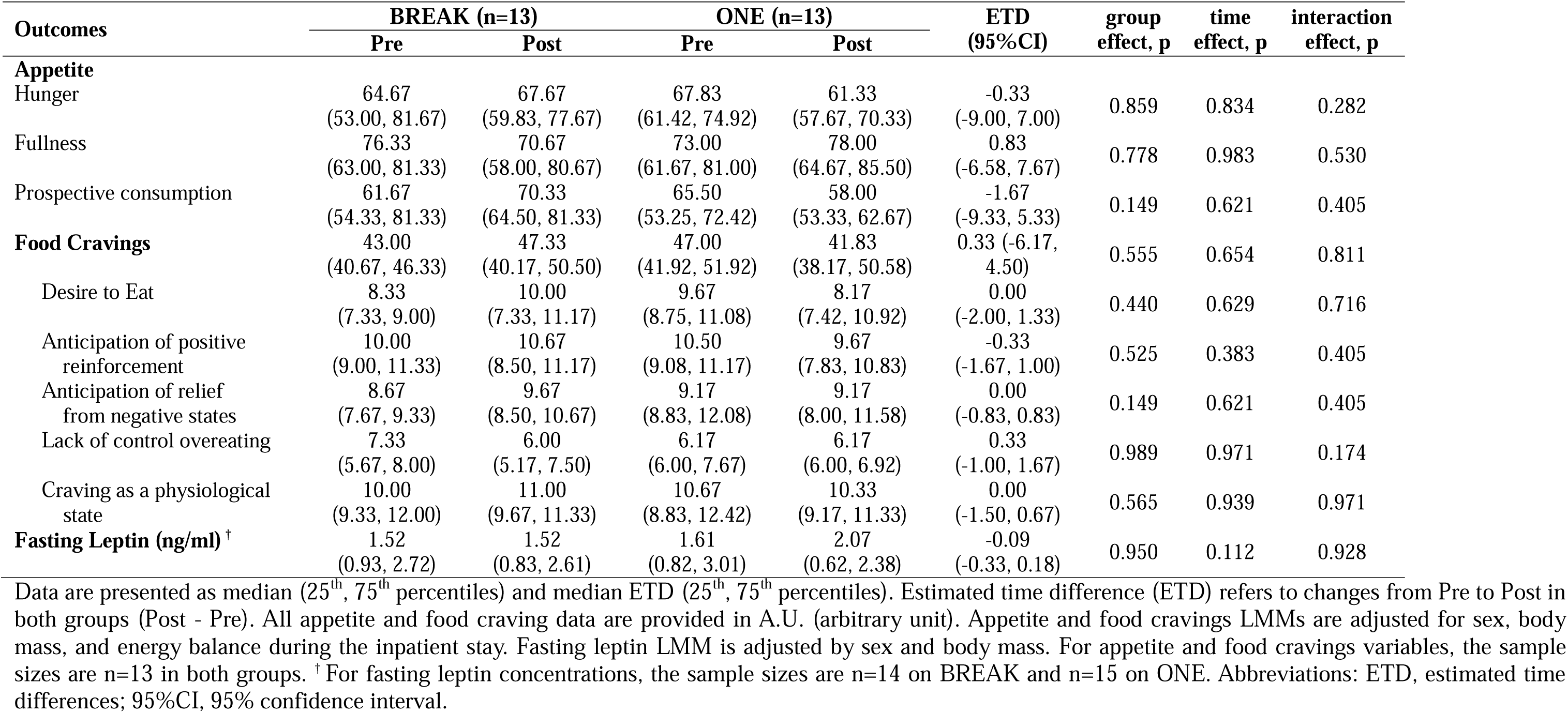
Appetite, food cravings, and fasting leptin.

## DISCUSSION

This is the first RCT to prospectively examine, over several weeks, whether breaking up SB with brief bouts of PA alters daily movement patterns and energy balance differently than a traditional single daily PA, matched for intensity and total duration, in sedentary adults with overweight or obesity. Main findings are (i) both interventions increased daily MVPA, step count and TDEE to a similar extent; (ii) only the BREAK intervention reduced time spent in prolonged SB (≥60 min); and (iii) BM and FM remained stable in BREAK, whereas a small increase was observed in ONE, suggesting that the pattern of PA accumulation may influence short-term energy balance regulation; however, these differences were modest.

Many studies have examined the acute and short-term effects of short active breaks, but few have tested such interventions over several weeks in free-living conditions (7, 31). Here, BREAK reduced the proportion of daily SB accumulated in prolonged bouts from 34.3 % to 26.5%, whereas no change was observed in ONE. This distinction is important because prolonged SB has been associated with less favorable cardiometabolic profiles independent of total sedentary time and MVPA (6, 32). However, although epidemiological studies support the relevance of SB patterns, the dose-response relationship for clinically meaningful benefits remains unknown. Therefore, the reduction in prolonged SB should be viewed as a favorable behavior change, but its clinical significance remains to be established.

Prescribing brisk walking for 45 min/day, 5 days/week, as a single PA bout or as repeated short bouts, increased daily MVPA in sedentary adults with overweight/obesity. Participants in both groups reached MVPA levels (BREAK: 38.8 min/day; ONE: 31.3 min/day) consistent with guidelines (>30 min/day) (33) and increased step count toward levels associated with lower chronic disease and mortality risk (8,000-10,000 steps/day) (34). This supports the broader public health message: “every movement counts” (33). Notably, both interventions also reduced SB by 30 min/day, a magnitude relevant to population health (35).

An important observation is that the prescribed MVPA did not simply replace SB. In both groups, increases in MVPA occurred at the expense of sedentary time and LPA, supporting the interdependence of daily movement components. A spontaneous reduction in non-exercise PA (e.g., daily walking, LPA) and increased sedentary time has been previously reported following exercise (MVPA) training, particularly in sedentary individuals with overweight/obesity (17, 36). Yet, contrary to our hypothesis (37), spreading PA as short bouts throughout the day did not better preserve LPA than a single continuous session matched for volume and intensity. Thus, while BREAK altered the pattern of sedentary accumulation, it did not produce a clearly different redistribution of overall daily movement composition than ONE. Nonetheless, population data show that replacing LPA with MVPA confers health benefits (38), including reductions in BM or adiposity (39), suggesting both interventions may improve health.

Like the behavioral data, the energetics data do not support the idea that BREAK reduced overall compensatory responses (decreases in RMR and/or lower-than-expected increases in AEE due to spontaneous behavioral adaptations) regularly reported in response to structured MVPA interventions (36, 40). As described by King et al., these compensatory responses may be driven by physiological fatigue, motivational factors (e.g., perceiving inactivity as a reward), and neuroregulatory processes protecting against sustained negative energy balance (41). Despite a modest reduction in LPA, both interventions increased TDEE to a similar extent, with no evidence of differential effects on AEE or RMR.

Estimated EI did not differ significantly between groups, although values were numerically higher in ONE than BREAK. Likewise, no changes in appetite ratings, food cravings, or fasting leptin were detected. These findings differ from our previous acute study, in which frequent short bouts of moderate-intensity PA reduced pre-meal food cravings, whereas a matched continuous bout did not, both compared to a sedentary condition (10). Several factors may explain this discrepancy, including differences in study design (acute 6-hour vs. 6-week), population (individuals with normal weight vs. overweight/obesity), and the fact that appetite was not assessed on days when the intervention itself was performed. It is also possible that appetite responses to altered movement patterns are subtle and influenced by non-homeostatic drivers of intake (e.g., food reward, palatability, environmental cues) that promote intake beyond homeostatic needs (42, 43) and were not captured here. More broadly, individuals with overweight or obesity often exhibit altered appetite regulation and may require stronger or longer perturbations to elicit measurable changes in appetite and food intake.

The small increase in BM and FM observed in ONE, but not in BREAK, is intriguing, as it was not clearly explained by differences in measured EI or expenditure. This may reflect a small cumulative energy imbalance not detected with the measures used (i.e., no pre-/post-DXA data, missing data, only a few weeks), or day-to-day variability that becomes relevant over longer time periods. However, these findings should be interpreted cautiously, as interaction terms were attenuated in sensitivity analyses, excluding the over-compliant BREAK participant, though likely reflecting reduced between-group contrast rather than reversal of changes in ONE. Taken together, these data highlight the need for future studies investigating the influence of PA accumulation patterns on long-term energy balance and body composition, and whether short, frequent bouts of PA differentially influence the compensatory adaptations often observed with exercise (MVPA) training.

Importantly, this study should be considered a proof-of-concept conducted under relatively controlled free-living conditions. Frequent activity breaks were supported by regular contact and motivational prompts, and the prescribed PA dose (45 min/day, 5 days/week) exceeds minimum recommendations. Therefore, the feasibility and effectiveness of less structured, scalable interventions remain to be established. Larger or more sustained changes in movement behavior may be required to elicit clinically meaningful health benefits.

This study has several strengths, including high adherence, direct PA/SB assessment using complementary devices (ActiGraph, ActivPAL), gold-standard measurement of free-living TDEE using DLW, and balanced sex distribution. Limitations include the relatively short duration, which may have limited detection of robust changes in body composition and energy balance components; the use of isotopic dilution rather than repeated DXA, although baseline agreement was strong; and the lack of port-prandial assessment of appetite-regulating hormones (e.g., ghrelin, GLP-1, PYY), which could provide insights given their links to energy compensation and FM loss (40, 44). The modest sample size for energetics outcome, due to technical issues, reduced power to detect small between-group differences.

## CONCLUSION

Both a single, daily bout of brisk walking and frequent short bouts accumulated across the day increased MVPA and TDEE in sedentary adults with overweight or obesity. Only BREAK reduced prolonged sedentary time. BM and FM remained stable in BREAK but increased modestly in ONE; however, these findings should be interpreted cautiously. Overall, breaking up sedentary time with short brisk walking bouts may improve daily movement patterns without compromising TDEE. Longer-term studies are warranted to determine whether these behavioral changes translate into clinically meaningful benefits for health.

## Supporting information

CONSORT checklist

## Data Availability

The trial protocol, statistical analysis plan, and deidentified datasets generated and analyzed during the current study are available from the corresponding author on reasonable request, via secure file transfer. Data can be shared with researchers for purposes of reproducing the results or conducting meta-analyses and will be available indefinitely from the time of publication.

## ACKOWLEDMENTS

The authors thank study participants for their time and commitment, as well as the medical, nutrition, and administrative staff of the University of Colorado Clinical and Translational Research Center (CTRC).

## REFERENCES

1. Pinto AJ, Bergouignan A, Dempsey PC, et al. Physiology of sedentary behavior. Physiol Rev 2023;103:2561–2622.

2. Rezende LFM, Ahmadi M, Ferrari G, et al. Device-measured sedentary time and intensity-specific physical activity in relation to all-cause and cardiovascular disease mortality: the UK Biobank cohort study. International Journal of Behavioral Nutrition and Physical Activity 2024;21:1–10.

3. Matthews CE, Keadle SK, Troiano RP, et al. Accelerometer-measured dose-response for physical activity, sedentary time, and mortality in US adults. American Journal of Clinical Nutrition 2016;104:1424–1432.

4. Liao J, Cao C, Hur J, et al. Association of sedentary patterns with body fat distribution among US children and adolescents: a population-based study. Int J Obes 2021;45:2048–2057.

5. Barone Gibbs B, Pettee Gabriel K, Carnethon MR, et al. Sedentary Time, Physical Activity, and Adiposity: Cross-sectional and Longitudinal Associations in CARDIA. Am J Prev Med 2017;53:764–771.

6. Healy GN, Dunstan DW, Salmon J, et al. Breaks in Sedentary: Time beneficial associations with metabolic risk. Diabetes Care 2008;31:661–666.

7. Hadgraft NT, Winkler E, Climie RE, et al. Effects of sedentary behaviour interventions on biomarkers of cardiometabolic risk in adults: systematic review with meta-analyses. Br J Sports Med 2020;55:144.

8. Swartz AM, Squires L, Strath SJ. Energy expenditure of interruptions to sedentary behavior. International Journal of Behavioral Nutrition and Physical Activity 2011;8:1–7.

9. Carter SE, Jones M, Gladwell VF. Energy expenditure and heart rate response to breaking up sedentary time with three different physical activity interventions. Nutr Metab Cardiovasc Dis 2015;25:503–509.

10. Bergouignan A, Legget KT, De Jong N, et al. Effect of frequent interruptions of prolonged sitting on self-perceived levels of energy, mood, food cravings and cognitive function. Int J Behav Nutr Phys Act 2016;13.

11. Dutta N, Koepp GA, Stovitz SD, Levine JA, Pereira MA. Using Sit-Stand Workstations to Decrease Sedentary Time in Office Workers: A Randomized Crossover Trial. Int J Environ Res Public Health 2014;11:6653.

12. World Medical Association - WMA. Declaration of Helsinki – Ethical Principles for Medical Research Involving Human Subjects. [WWW document]. URL https://www.wma.net/policies-post/wma-declaration-of-helsinki-ethical-principles-for-medical-research-involving-human-subjects/

13. De Jong NP, Rynders CA, Goldstrohm DA, et al. Effect of frequent interruptions of sedentary time on nutrient metabolism in sedentary overweight male and female adults. J Appl Physiol 2019;126:984–992.

14. Edwardson CL, Winkler EAH, Bodicoat DH, et al. Considerations when using the activPAL monitor in field-based research with adult populations. J Sport Health Sci 2017;6:162.

15. Trost SG, Mciver KL, Pate RR. Conducting accelerometer-based activity assessments in field-based research. Med Sci Sports Exerc 2005;37.

16. Freedson PS, Melanson E, Sirard J. Calibration of the Computer Science and Applications, Inc. accelerometer. Med Sci Sports Exerc 1998;30:777–781.

17. Lefai E, Blanc S, Momken I, et al. Exercise training improves fat metabolism independent of total energy expenditure in sedentary overweight men, but does not restore lean metabolic phenotype. Int J Obes 2017;41:1728–1736.

18. Heymsfield SB, Wang ZM, Baumgartner RN, Ross R. Human body composition: Advances in models and methods. Annu Rev Nutr 1997;17:527–558.

19. Sheng HP, Huggins RA. A review of body composition studies with emphasis on total body water and fat. Am J Clin Nutr 1979;32:630–647.

20. Bergouignan A, Momken I, Schoeller DA, et al. Regulation of energy balance during long-term physical inactivity induced by bed rest with and without exercise training. Journal of Clinical Endocrinology and Metabolism 2010;95:1045–1053.

21. Schoeller DA, Ravussin E, Schutz Y, Acheson KJ, Baertschi P, Jéquier E. Energy expenditure by doubly labeled water: validation in humans and proposed calculation. Am J Physiol 1986;250.

22. Melanson EL, Ingebrigtsen JP, Bergouignan A, Ohkawara K, Kohrt WM, Lighton JRB. A new approach for flow-through respirometry measurements in humans. Am J Physiol Regul Integr Comp Physiol 2010;298.

23. Villalon KL, Gozansky WS, Van Pelt RE, et al. A losing battle: Weight regain does not restore weight loss-induced bone loss in postmenopausal women. Obesity 2011;19:2345–2350.

24. Racette SB, Das SK, Bhapkar M, et al. Approaches for quantifying energy intake and %calorie restriction during calorie restriction interventions in humans: the multicenter CALERIE study. Am J Physiol Endocrinol Metab 2012;302.

25. Hill JO, Wyatt HR, Peters JC. Energy Balance and Obesity. Circulation 2012;126:126.

26. Thomas DM, Martin CK, Heymsfield S, Redman LM, Schoeller DA, Levine JA. A Simple Model Predicting Individual Weight Change in Humans. J Biol Dyn 2011;5:579.

27. Dempsey PC, Larsen RN, Sethi P, et al. Benefits for Type 2 Diabetes of Interrupting Prolonged Sitting With Brief Bouts of Light Walking or Simple Resistance Activities. Diabetes Care 2016;39:964–972.

28. Meule A. Twenty Years of the Food Cravings Questionnaires: a Comprehensive Review. Current Addiction Reports 2020 7:1 2020;7:30–43.

29. Cepeda-Benito A, Gleaves DH, Williams TL, Erath SA. The development and validation of the state and trait food-cravings questionnaires. Behav Ther 2000;31:151–173.

30. Chastin SFM, Palarea-Albaladejo J, Dontje ML, Skelton DA. Combined Effects of Time Spent in Physical Activity, Sedentary Behaviors and Sleep on Obesity and Cardio-Metabolic Health Markers: A Novel Compositional Data Analysis Approach. PLoS One 2015;10:e0139984.

31. Buffey AJ, Herring MP, Langley CK, Donnelly AE, Carson BP. The Acute Effects of Interrupting Prolonged Sitting Time in Adults with Standing and Light-Intensity Walking on Biomarkers of Cardiometabolic Health in Adults: A Systematic Review and Meta-analysis. Sports Medicine 2022 52:8 2022;52:1765–1787.

32. Healy GN, Matthews CE, Dunstan DW, Winkler EAH, Owen N. Sedentary time and cardio-metabolic biomarkers in US adults: NHANES 2003–06. Eur Heart J 2011;32:590.

33. Bull FC, Al-Ansari SS, Biddle S, et al. World Health Organization 2020 guidelines on physical activity and sedentary behaviour. Br J Sports Med 2020;54:1451.

34. Ding D, Nguyen B, Nau T, et al. Daily steps and health outcomes in adults: a systematic review and dose-response meta-analysis. Lancet Public Health 2025;10:e668–e681.

35. Boudreaux BD, Xu C, Serafini MA, et al. Achieving Guidelines Within a 24-Hour Movement Paradigm and Risk of Mortality in US Adults. Mayo Clin Proc 2026.

36. Riou MÈ, Jomphe-Tremblay S, Lamothe G, et al. Energy Compensation Following a Supervised Exercise Intervention in Women Living With Overweight/Obesity Is Accompanied by an Early and Sustained Decrease in Non-structured Physical Activity. Front Physiol 2019;10.

37. Bourdier P, Simon C, Bessesen DH, Blanc S, Bergouignan A. The role of physical activity in the regulation of body weight: The overlooked contribution of light physical activity and sedentary behaviors. Obesity Reviews 2023;24:e13528.

38. Miatke A, Olds T, Maher C, et al. The association between reallocations of time and health using compositional data analysis: a systematic scoping review with an interactive data exploration interface. Int J Behav Nutr Phys Act 2023;20.

39. Grgic J, Dumuid D, Bengoechea EG, et al. Health outcomes associated with reallocations of time between sleep, sedentary behaviour, and physical activity: a systematic scoping review of isotemporal substitution studies. Int J Behav Nutr Phys Act 2018;15.

40. Flack KD, Hays HM, Moreland J, Long DE. Exercise for Weight Loss: Further Evaluating Energy Compensation with Exercise. Med Sci Sports Exerc 2020;52:2466.

41. King NA, Caudwell P, Hopkins M, et al. Metabolic and Behavioral Compensatory Responses to Exercise Interventions: Barriers to Weight Loss. Obesity 2007;15:1373–1383.

42. Lowe MR, Butryn ML. Hedonic hunger: A new dimension of appetite? Physiol Behav 2007;91:432–439.

43. Blundell JE, Finlayson G. Is susceptibility to weight gain characterized by homeostatic or hedonic risk factors for overconsumption? Physiol Behav 2004;82:21–25.

44. Jin Z, Li J, Thackray AE, et al. Fasting appetite-related gut hormone responses after weight loss induced by calorie restriction, exercise, or both in people with overweight or obesity: a meta analysis: Physiology and Biochemistry. Int J Obes 2025;49:776–792.

