## Supplementary File for "Frequent vs single active bouts differentially affect movement behavior and energy balance in adults with overweight/obesity"

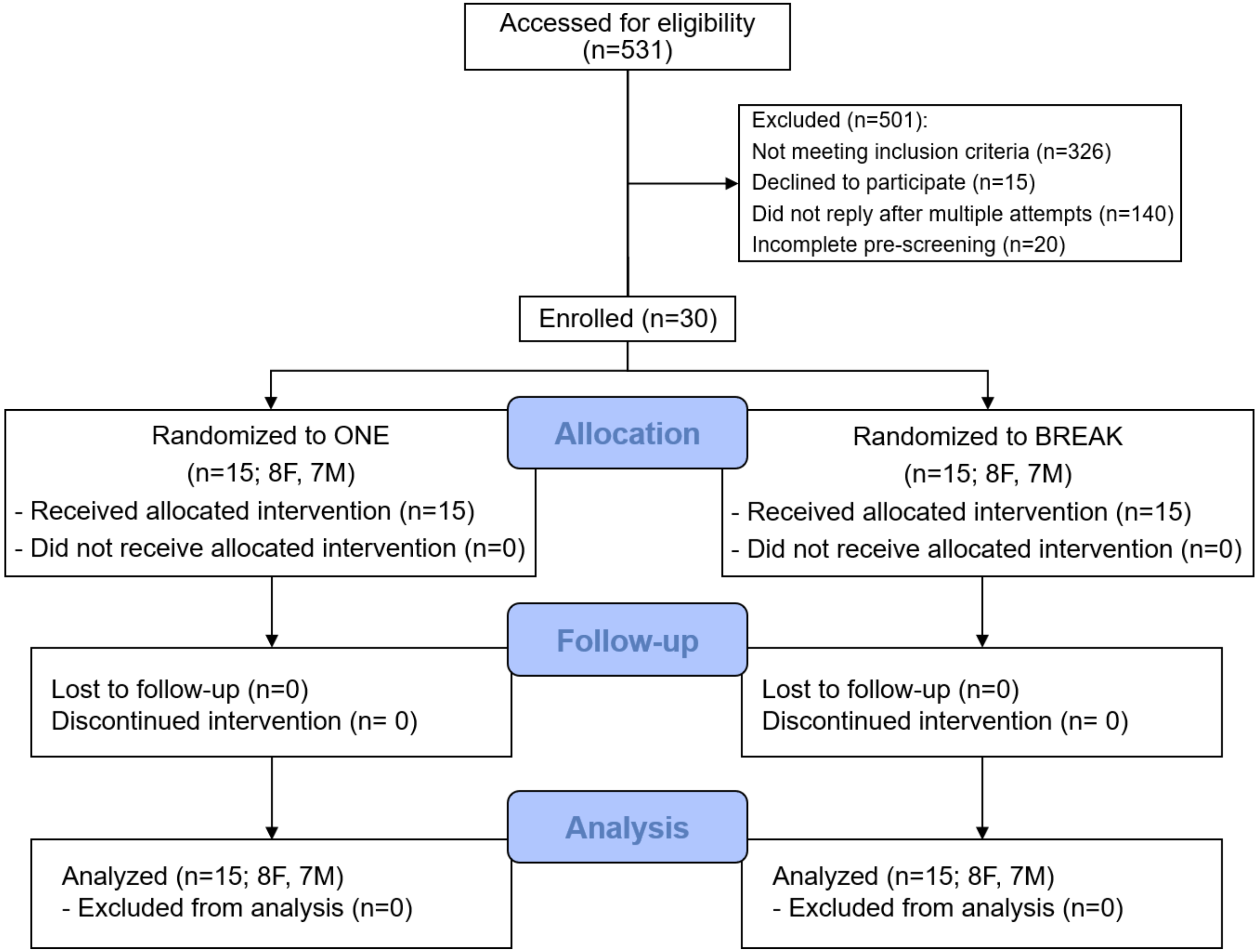


**Supplementary Figure 1.** Consolidated Standards of Reporting Trials (CONSORT) flow diagram. Exclusion and missing data for specific outcomes are described in Supplementary Table 2.Abbreviations: BREAK, 9 hourly 5-min brisk walking bouts, 5 d/wk; ONE, single 45-min brisk walking bout, 5 d/wk.

**Supplementary Table 1.** Participants with complete data and reasons for missing data for all study outcomes.

| Outcomes | Sample size | | | | Reasons |
|---|---|---|---|---|---|
| | Break - Pre | Break - Post | One - Pre | One - Post | |
| Sedentary behavior and physical activity (ActiGraph) | 15 | 14 | 14 | 13 | Faulty or lost device. |
| Sedentary behavior and physical activity (ActivPAL) | 15 | 15 | 15 | 14 | Lost device. |
| Total daily energy expenditure, activity energy expenditure, and physical activity level | 11 | 13 | 12 | 14 | All quality checks were not met when subjects were dosed with one out of the six doubly labelled water doses; and one participant did not return the samples back. |
| Resting metabolic rate | 15 | 15 | 15 | 15 | Not applicable. |
| Body composition | 10 | 12 | 11 | 12 | All quality checks were not met when subjects were dosed with one out of six doubly labelled water doses; and one participant did not return the samples back. |
| Metabolized energy intake | - | 10 | - | 9 | Outcome derived from body composition and total daily energy expenditure in Pre and Post. |
| Perceived appetite and food cravings | 13 | 12 | 13 | 13 | Incomplete data for one or more meals. |
| Fasting leptin concentrations | 14 | 13 | 15 | 15 | Out of range values. |

**Supplementary Table 2.** Model estimates for isometric log-ratio (ilr) coordinates of the movement compositions.

| Outcomes | BREAK (n=15) | | ONE (n=15) | | ETD (95%CI) | EMD (95%CI) | group effect, p | time effect, p | interaction effect, p |
|---|---|---|---|---|---|---|---|---|---|
| | Pre | Post | Pre | Post | | | | | |
| **ActivPAL** | | | | | | | | | |
| Total SB | | | | | | | | | |
| ilr1 (SB vs standing+stepping) | 1.33 (1.12, 1.54) | 1.15 (0.94, 1.36) | 1.25 (1.04, 1.45) | 1.04 (0.83, 1.25) | -0.20 (-0.34, -0.05) | 0.03 (-0.26, 0.32) | 0.453 | 0.011 | 0.842 |
| ilr2 (standing vs stepping) | 0.67 (0.57, 0.76) | 0.47 (0.37, 0.56) | 0.53 (0.43, 0.62) | 0.42 (0.32, 0.51) | -0.15 (-0.23, -0.08) | -0.08 (-0.23, 0.06) | 0.113 | <0.001 | 0.252 |
| Standing | | | | | | | | | |
| ilr1 (standing vs SB+stepping) | -0.09 (-0.23, 0.05) | -0.17 (-0.31, -0.03) | -0.17 (-0.30, -0.03) | -0.16 (-0.30, -0.02) | -0.04 (-0.13, 0.05) | -0.08 (-0.25, 0.09) | 0.678 | 0.357 | 0.341 |
| ilr2 (SB vs stepping) | 1.48 (1.30, 1.67) | 1.23 (1.05, 1.41) | 1.34 (1.16, 1.53) | 1.10 (0.92, 1.29) | -0.25 (-0.39, -0.11) | -0.02 (-0.29, 0.26) | 0.241 | 0.001 | 0.912 |
| Stepping | | | | | | | | | |
| ilr1 (stepping vs SB+standing) | -1.24 (-1.37, -1.11) | -0.98 (-1.11, -0.85) | -1.08 (-1.21, -0.95) | -0.88 (-1.01, -0.74) | 0.23 (0.13, 0.34) | 0.06 (-0.15, 0.27) | 0.096 | <0.001 | 0.581 |
| ilr2 (SB vs standing) | 0.82 (0.63, 1.01) | 0.76 (0.57, 0.95) | 0.82 (0.63, 1.00) | 0.69 (0.50, 0.89) | -0.09 (-0.21, 0.03) | 0.06 (-0.19, 0.31) | 0.770 | 0.149 | 0.606 |
| **ActiGraph** | | | | | | | | | |
| Inactivity | | | | | | | | | |
| ilr1 (inactivity vs LPA+MVPA) | 1.90 (1.66, 2.13) | 1.53 (1.29, 1.77) | 1.78 (1.54, 2.02) | 1.53 (1.28, 1.78) | -0.31 (-0.53, -0.09) | -0.11 (-0.55, 0.33) | 0.669 | 0.007 | 0.600 |
| ilr2 (LPA vs MVPA) | 1.83 (1.54, 2.12) | 1.18 (0.88, 1.48) | 1.96 (1.65, 2.26) | 1.44 (1.13, 1.75) | -0.58 (-0.85, -0.31) | -0.13 (-0.67, 0.41) | 0.251 | <0.001 | 0.621 |
| LPA | | | | | | | | | |
| ilr1 (LPA vs inactivity+MVPA) | 0.63 (0.43, 0.84) | 0.26 (0.05, 0.47) | 0.82 (0.61, 1.02) | 0.48 (0.26, 0.69) | -0.36 (-0.52, -0.19) | -0.04 (-0.36, 0.28) | 0.117 | <0.001 | 0.803 |
| ilr2 (inactivity vs MVPA) | 2.56 (2.24, 2.87) | 1.91 (1.59, 2.24) | 2.52 (2.20, 2.85) | 2.05 (1.71, 2.38) | -0.56 (-0.86, -0.25) | -0.16 (-0.77, 0.45) | 0.780 | 0.001 | 0.586 |
| MVPA | | | | | | | | | |
| ilr1 (MVPA vs inactivity+LPA) | -2.53 (-2.87, -2.19) | -1.79 (-2.13, -1.44) | -2.58 (-2.93, -2.24) | -2.01 (-2.37, -1.65) | 0.66 (0.33, 0.98) | 0.17 (-0.48, 0.82) | 0.461 | <0.001 | 0.589 |
| ilr2 (inactivity vs LPA) | 0.73 (0.57, 0.89) | 0.73 (0.56, 0.90) | 0.54 (0.38, 0.71) | 0.60 (0.43, 0.78) | 0.03 (-0.08, 0.14) | -0.06 (-0.10, 0.23) | 0.146 | 0.584 | 0.591 |

† Data are presented as LSMeans (95%CI), ETD (95%CI), and EMD (95%CI), calculated by LMMs. ETD refers to changes from Pre to Post in both groups (Post - Pre), whereas EMD refers to differences between changes in BREAK and in ONE (ΔBREAK - ΔONE). Positive ETD values indicate an increase in the first behavior of the ilr coordinate relative to the other behaviors over time, whereas positive EMD values indicate a greater increase in the first behavior of the ilr coordinate in BREAK compared with ONE. LMMs were adjusted by sex. Sensitivity analyses yielded similar conclusions. Abbreviations: ETD, estimated time differences; EMD, estimated mean difference; ilr, isometric log-ratio; LPA, light physical activity; MVPA, moderate-to-vigorous physical activity; SB, sedentary behavior; 95%CI, 95% confidence interval.

**Supplementary Table 3.** Sex-adjusted model estimates for energy expenditure, appetite, food cravings, and fasting leptin.

| Outcomes | Break (n=15) | | One (n=15) | | ETD (95%CI) | EMD (95%CI) | group effect, p | time effect, p | interaction effect, p |
|---|---|---|---|---|---|---|---|---|---|
| | Pre | Post | Pre | Post | | | | | |
| **Energy expenditure** | | | | | | | | | |
| Total energy daily expenditure | 10.03 (8.86-11.20) | 10.51 (9.39-11.63) | 11.01 (9.89-12.13) | 12.01 (10.94-13.09) | 0.74 (0.08-1.40) | -0.53 (-1.86-0.79) | 0.094 | 0.030 | 0.412 |
| Resting metabolic rate | 6.67 (6.18-7.15) | 6.79 (6.30-7.28) | 6.71 (6.22-7.20) | 6.86 (6.38-7.35) | 0.14 (-0.02-0.29) | -0.03 (-0.34-0.28) | 0.857 | 0.077 | 0.851 |
| Activity energy expenditure | 2.27 (1.53-3.01) | 2.56 (1.88-3.25) | 3.25 (2.54-3.95) | 4.01 (3.35-4.67) | 0.53 (-0.02-1.08) | -0.47 (-1.57-0.63) | 0.007 | 0.058 | 0.383 |
| **Appetite** | | | | | | | | | |
| Hunger | 64.67 (53.00, 81.67) | 67.67 (59.83, 77.67) | 67.83 (61.42, 74.92) | 61.33 (57.67, 70.33) | -0.33 (-9.00, 7.00) | - | 0.573 | 0.781 | 0.347 |
| Fullness | 76.33 (63.00, 81.33) | 70.67 (58.00, 80.67) | 73.00 (61.67, 81.00) | 78.00 (64.67, 85.50) | 0.83 (-6.58, 7.67) | - | 0.837 | 0.916 | 0.715 |
| Prospective consumption | 61.67 (54.33, 81.33) | 70.33 (64.50, 81.33) | 65.50 (53.25, 72.42) | 58.00 (53.33, 62.67) | -1.67 (-9.33, 5.33) | - | 0.107 | 0.895 | 0.118 |
| **Food Craving** | 43.00 (40.67, 46.33) | 47.33 (40.17, 50.50) | 47.00 (41.92, 51.92) | 41.83 (38.17, 50.58) | 0.33 (-6.17, 4.50) | - | 0.822 | 0.609 | 0.831 |
| Desire to Eat | 8.33 (7.33, 9.00) | 10.00 (7.33, 11.17) | 9.67 (8.75, 11.08) | 8.17 (7.42, 10.92) | 0.00 (-2.00, 1.33) | - | 0.671 | 0.612 | 0.730 |
| Anticipation of positive reinforcement | 10.00 (9.00, 11.33) | 10.67 (8.50, 11.17) | 10.50 (9.08, 11.17) | 9.67 (7.83, 10.83) | -0.33 (-1.67, 1.00) | - | 0.413 | 0.417 | 0.390 |
| Anticipation of relief from negative states | 8.67 (7.67, 9.33) | 9.67 (8.50, 10.67) | 9.17 (8.83, 12.08) | 9.17 (8.00, 11.58) | 0.00 (-0.83, 0.83) | - | 0.327 | 0.430 | 0.462 |
| Lack of control overeating | 7.33 (5.67, 8.00) | 6.00 (5.17, 7.50) | 6.17 (6.00, 7.67) | 6.17 (6.00, 6.92) | 0.33 (-1.00, 1.67) | - | 0.910 | 0.972 | 0.171 |
| Craving as a physiological state | 10.00 (9.33, 12.00) | 11.00 (9.67, 11.33) | 10.67 (8.83, 12.42) | 10.33 (9.17, 11.33) | 0.00 (-1.50, 0.67) | - | 0.868 | 0.767 | 0.574 |
| **Fating Leptin** | 1.52 (0.93, 2.72) | 1.52 (0.83, 2.61) | 1.61 (0.82, 3.01) | 2.07 (0.62, 2.38) | -0.09 (-0.33, 0.18) | - | 0.905 | 0.127 | 0.971 |

Energy expenditure data are presented as LSMeans (95%CI), ETD (95%CI), and EMD (95%CI). Appetite, food craving and leptin data are presented as median (25th, 75th percentiles) and median ETD (25th, 75th percentiles). Abbreviations: ETD, estimated time differences; EMD, estimated mean difference; 95%CI, 95% confidence interval. Sensitivity analyses excluding adjustment by sex lead to similar conclusions.
